# Predictors of Response to Cardiac Resynchronization Therapy in Pediatric and Congenital Heart Disease Patients

**DOI:** 10.1101/2024.12.12.24318957

**Authors:** Henry Chubb, Douglas Mah, Maully Shah, Kimberly Y Lin, David Peng, Benjamin W Hale, Lindsay May, Susan Etheridge, William Goodyer, Scott R Ceresnak, Kara S Motonaga, David N Rosenthal, Christopher S Almond, Doff B McElhinney, Anne M Dubin

**Affiliations:** Division of Pediatric Cardiology, Department of Pediatrics, Stanford University, CA, USA; Division of Pediatric Cardiothoracic Surgery, Department of Cardiothoracic Surgery, Stanford University, CA, USA; Department of Cardiology, Boston Children’s Hospital; Department of Pediatrics, Harvard Medical School; Boston MA, USA; Department of Cardiology, Children’s Hospital of Philadelphia, Philadelphia, PA, USA; Department of Cardiology, CS Mott Children’s Hospital, University of Michigan, Ann Arbor, MI, USA; Department of Pediatrics, Stead Family Children’s Hospital, Iowa City, IA; Department of Cardiology, Primary Children’s Hospital, University of Utah, Salt Lake City, UT, USA

**Keywords:** Cardiac Resynchronization Therapy, Pediatric, Heart Failure, Congenital Heart Disease, Cardiac Transplant, Electrical Dyssynchrony, Cardiac Failure, Pediatric, Predictors of Response, Dyssynchrony

## Abstract

**Background:** Cardiac resynchronization therapy (CRT) is an important therapeutic option in selected pediatric and congenital heart disease (CHD) patients with reduced systemic ventricular (SV) ejection fraction (EF). However, the identification of optimal responders is challenging.

**Objective:** To identify predictors of response to CRT in children and CHD patients at 5 large quaternary referral centers.

**Methods:** Patients were <21 years or had CHD; had SVEF <45%; symptomatic heart failure; and significant electrical dyssynchrony prior to CRT. Primary outcome was defined as an ordinal response at 6/12months: (1) Improved EF [≥5%], (2) Unchanged SVEF, (3) Worse SVEF. Secondary outcome utilized a propensity score-matched control cohort. Response to CRT was defined using longitudinal trajectory of SVEF up to latest follow-up.

**Results:** In total, 167 eligible CRT recipients were identified across the 5 centers. 150 had comprehensive data at 6/12months: 96(64%) with improved SVEF, 26(17%) unchanged, 28(19%) worsened. Mean increase in SVEF was 11% [IQR 3-21%]. On univariable ordinal regression, lower SVEF (p=0.013), biventricular circulation (p=0.022), systemic LV (p=0.021), and conduction delay to lateral wall of SV (p=0.01) were associated with positive response.

For assessment of secondary outcome, 324 controls were identified. Mean follow-up 4.2(±3.7) yrs. Almost all subgroups demonstrated improved SVEF trend with CRT, except those with systemic RV (p=0.69) or without prior single site pacemaker (p=0.20).

**Conclusion:** CRT in children and CHD patients frequently results in an improvement in SVEF. Those with lower SVEF, conduction delay to lateral wall of the SV and those with systemic LV are most likely to respond.

**Condensed Abstract:** Cardiac resynchronization therapy (CRT) is an important therapeutic option in selected pediatric and congenital heart disease (CHD) patients with reduced systemic ventricular (SV) ejection fraction (EF). However, the identification of optimal responders is challenging.

In this multicenter study, pediatric and CHD CRT recipients with heart failure and EF<45% were identified. The primary outcome was change in EF at 6/12months. Those with lower baseline SVEF, conduction delay to lateral wall of the SV and/or systemic LV were most likely to respond to CRT. When compared to propensity score-matched controls, the CRT groups also demonstrated a significantly better long-term EF trajectory.

## Introduction

Cardiac resynchronization therapy (CRT) is a treatment modality widely employed for patients with significant systemic ventricular (SV) dysfunction and evidence of electrical dyssynchrony. However, evidence for the use of this important treatment is relatively limited in children and congenital heart disease (CHD) patients as all prospective randomized trials of CRT have specifically excluded these subjects.^1–6^ None the less, retrospective studies and meta-analysis have clearly demonstrated that many children and CHD patients may benefit from CRT in terms of improvement in systemic ventricular function.^7–11^ Furthermore, we have recently demonstrated in a multicenter study that there is a strong association of CRT with survival in a broad group of patients with systemic ventricular ejection fraction (SVEF)<45%, significant electrical dyssynchrony and cardiac failure symptoms.^12^ However, the factors that identify those children and CHD subjects that are most likely to respond to CRT have not been defined.

The need to identify the pediatric and CHD groups that are most likely to respond to CRT is particularly pertinent in the context of the increased technical challenges of CRT implant procedures compared to adults with structurally normal hearts. The implants may be highly complex and frequently require repeat sternotomy. In adults without CHD, the presence of left bundle branch block (LBBB), female sex, and longer QRS duration (QRSd) have been identified to be the most robust predictors of response.^13^ In the pediatric/CHD patients, though, QRSd>150msec and LBBB are rarely seen.^14^

In a population as heterogenous as that of children and CHD subjects with systemic ventricular systolic failure, there are many fundamental pathophysiological differences between patients that are likely to play a role in response to CRT. However, the largest retrospective studies of these patients have not concurred as to whether there are distinct subgroups that are more or less likely to respond. Key subgroups for assessment include those with and without prior ventricular pacing, those with and without CHD and then, within CHD, those with and without a biventricular circulation and systemic LV.^11, 15^

We hypothesized that the assessment within a larger multicenter CRT cohort would be sufficiently powered to identify key predictors of response to CRT within these patient groups. Given the challenge in defining response in these heterogenous morphologies, we chose to use two main outcomes. The first was medium term (6-12month) change in SVEF, referenced to the patient baseline prior to CRT. The second utilized a matched control group (without CRT) to compare the longitudinal impact of CRT.

## Methods

### Data collection

This was a multicenter retrospective study. Patient data were de-identified and electronically transferred to the coordinating center through a secure web-based server (REDCap [Research Electronic Data Capture], Vanderbilt University, Nashville, TN). The study was approved by institutional review board (IRB) at the co-ordinating center (Stanford University, IRB-45389), with local IRB and data use agreement approval obtained at each center. The survival outcomes of this cohort have recently been reported.^12^

### Study population

All pediatric (<21years) and/or CHD patients who underwent CRT at five large quaternary referral institutions (Lucile Packard Children’s Hospital and Stanford Healthcare, Boston Children’s Hospital, Children’s Hospital of Philadelphia, Primary Children’s Hospital (University of Utah), and CS Mott Children’s Hospital (University of Michigan)) between Jan 1, 2004 and Jan 1, 2018 were identified. CRT was defined as a multi-site ventricular pacing system implanted with at least one pacing lead to the systemic ventricle (SV). CHD was defined as any congenital heart defect other than isolated bicuspid aortic valve, patent foramen ovale or patent ductus arteriosus. Subjects were included if, at the time of CRT implant, they had systemic SVEF <45%; symptomatic heart failure [defined as American Heart Association Stage C or D]; and significant electrical dyssynchrony [defined as either a QRSd z-score≥3 or, in paced patients, a ventricular pacing burden (Vp) ≥40%] (Figure 1). Subjects were excluded if they had Eisenmenger syndrome, a current ventricular assist device, previous heart transplant or weight <4kg.

**Figure 1.**
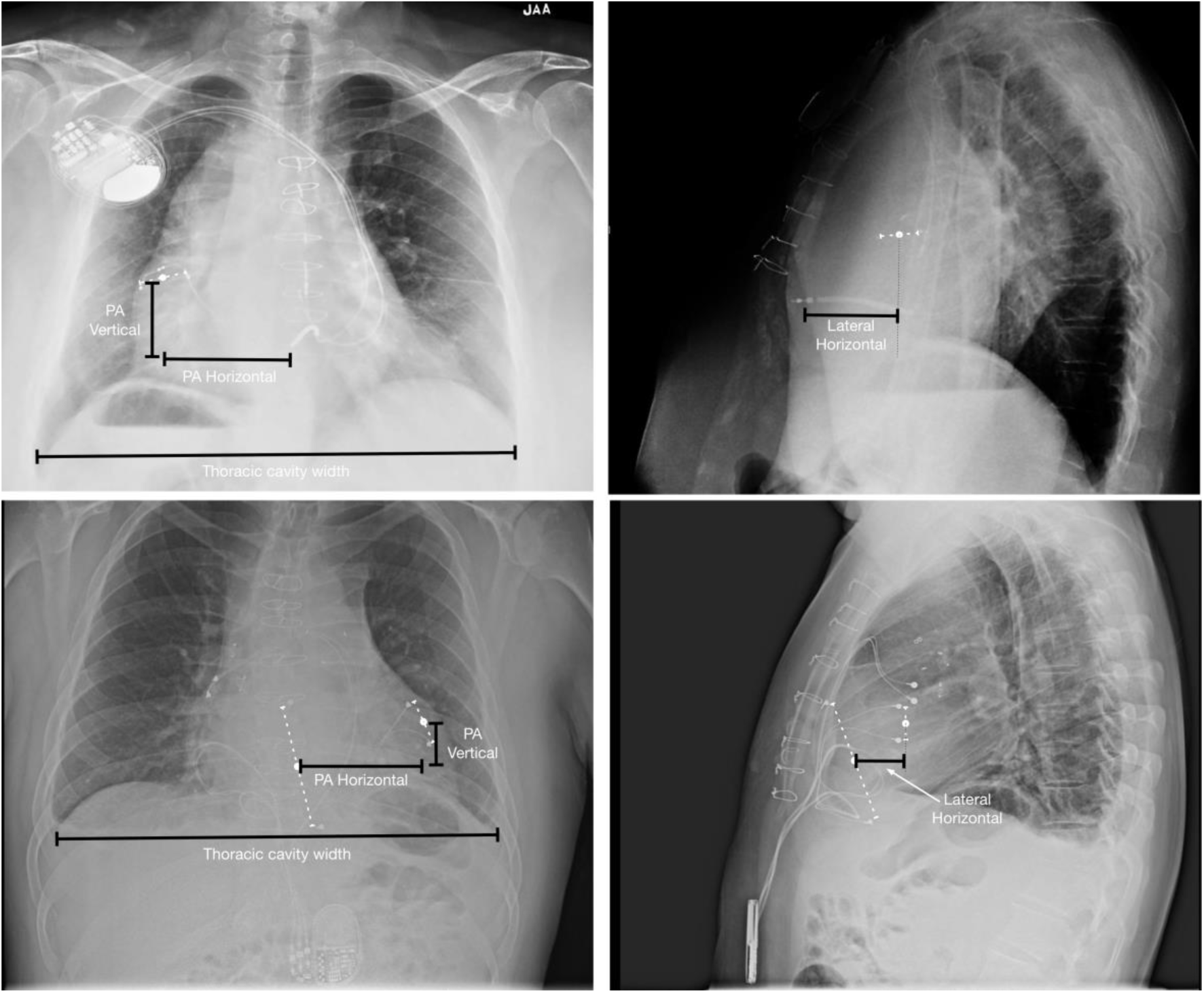
Measurement of separation of the ventricular leads. Top row-endovascular system, bottom row-epicardial system.

Control subjects were defined as pediatric and/or CHD patients who never received CRT and who met the same inclusion and exclusion criteria at an outpatient clinical encounter within the same time period. Controls were also excluded if there was a documented, planned, intervention for heart failure, such as alleviation of systemic outflow tract obstruction or ablation procedure for high ventricular ectopy burden. However, controls became eligible for enrollment once the intervention had been performed, and, similarly, once weight was >4kg, provided all inclusion criteria continued to be met.

### Identification of Subjects, Enrollment and Follow-Up

CRT subjects were identified via the institutional pacing databases. For control subjects, a comprehensive screening cohort with low SVEF and electrical dyssynchrony was identified by cross-referencing the institutional echocardiographic, ECG and pacing databases. All patients with both an estimated SVEF <45% and QRSd z-score ≥3 were selected for individual review for fulfillment of inclusion and exclusion criteria. Ventricular pacing-related dyssynchrony was identified via cross-referencing the echocardiographic database with the temporally matched device interrogation report to identify Vp >40% and SVEF <45%. Enrollment for CRT subjects was on the day of CRT implantation. Enrollment for control subjects was at the first outpatient appointment where all inclusion criteria were met.

Longitudinal assessments of all patients were collected at follow-up closest to 6 months, 1 year, and 5 years post enrollment, and latest follow-up. Precise date of assessment, number of heart failure medications, NYHA class, SVEF, QRSd, height and weight were obtained at each timepoint.

#### Electrical Assessment

QRSd was measured in leads II and V_5_ and averaged. Strict left bundle branch block (LBBB) was defined using adaptation of the criteria outlaid by Strauss et al, and was assessed on the surface ECG without reference to underlying cardiac anatomy.^16^ Right bundle branch block (RBBB) was defined according to the Minnesota Code criteria (7-2-1). QRSd z-score equivalent to ≥120ms (z-score ≥+2.3) was used along with the published RBBB criteria.^17^

Conduction delay to the lateral wall of the SV (CDtoLWSV) was defined as those patients with strict criteria LBBB and SV (which could be morphological left or right ventricle) in the conventional left ventricular (LV) position, *or* a RBBB and SV in conventional right ventricular (RV) position. Those with complex ventricular arrangement (such as dextrocardia) therefore could not be defined as CDtoLWSV, regardless of the ECG morphology.

#### Radiographic Assessment

Separation of the ventricular pacing leads was assessed on the first biplane chest x-ray following CRT implant (Figure 1). Unipolar pacing was rarely employed, and for those with multiple poles (eg epicardial bipolar leads), the mid-point of all poles was taken for each ventricular lead. Total distance in 3-dimensions, assuming orthogonal alignment of the two imaging planes, was indexed to the thoracic cavity width, creating a Lead Separation Ratio (LSR).

#### Echocardiographic Assessment

Baseline echocardiographic measurements were made on the final comprehensive echocardiogram prior to CRT implant. SVEF for the systemic RV was calculated from the fractional area change (RVFAC), using the estimation RVEF = 10.7 + (0.87*RVFAC).^18^

### First Definition of Response: Medium-Term Change in SVEF from Baseline

There is no clear consensus on the optimal marker of response to CRT, particularly in the CHD population where many patients have differing systemic ventricular morphology. Most non-CHD studies of CRT response have used a ≥15% reduction in LVESV as a threshold, but such a measure is not universally applicable in patients with mixed ventricular anatomy. ^13, 19^ Studies of CRT including CHD subjects have historically used a variety of measures of change in SV ejection fraction EF.^7–9, 11^ A primary outcome of an echocardiographic functional response was therefore defined as follows:

1. Worsened: fall in SVEF at follow-up following CRT implant
2. No change: absolute increase in SVEF of between 0% and 5%
3. Improvement: absolute increase in SVEF of ≥5%

Other studies have used a proportional increase in SVEF as a marker of response, with the rationale that larger (but sometimes less clinically significant) changes may occur in patients with higher baseline EF, potentially biasing results. Therefore, a proportional response was also defined in order to further enable comparison between studies:

1. Worsened: fall in SVEF at follow-up following CRT implant
2. No change: proportional increase in SVEF of between 0% and 10%
3. Improvement: proportional increase in SVEF of ≥10%

Echocardiographic parameters were assessed at closest follow-up to 6 months post implant and 12 months post implant for all patients. Follow-up at both time points was available in many, but not all, patients. Following sensitivity analyses for those with assessments recorded at both 6 and 12 months (see results), the average of the two measurements was used in order to mitigate for variations in time to assessment.

### Second Definition of Response: Longitudinal Trends with and without CRT

The identification of propensity score matched controls has been described in detail in a prior publication from our group,^12^ and the tables detailing the baseline parameters of the unmatched and matched cohorts meeting inclusion criteria for each analysis are included in the Data Supplement. Response to CRT was defined as a significant positive impact of CRT upon longitudinal modelling of the long-term parameters (see statistical details below).

### Statistics

Mean ± standard deviation was used to summarize normally distributed continuous variables, and median (with 25^th^-75^th^ percentiles) was used for non-normal distribution or non-continuous ordinal data. Categorical variables are presented as number (%). A priori, six anatomical and pathophysiological subgroups were established by firstly dividing the cohort into those with and without prior significant ventricular pacing (defined as Vp>40% prior to CRT), and then further subdividing into three anatomical subgroups: those without CHD, those with CHD with two ventricles and systemic LV, and those with other (‘complex’) CHD. Between-group variation was quantified using one-way ANOVA for normally distributed variables and Kruskall-Wallis for non-normal/categorical variables.

Ordinal logistic regression was used to assess the association of indices with ordinal outcomes (worse/no change/improved absolute or proportional change in SVEF). Multivariable modelling used a stepwise approach taking forward parameters with p<0.05 on univariable analysis.

Multivariable modelling was performed without inclusion of the subgroup 1-6 variable on account of collinearity, except on direct assessment of the significant subgroups. Longitudinal measures were evaluated using multilevel mixed-effects linear regression modeling with cubic model for estimation of pacing effect and visualized using fractional polynomial plot with 95% CIs. Statistics were analyzed using Stata (version 18.0, Stata Corp LP, College Station, Texas).

## Results

### Subjects

Baseline characteristics are delineated in Table 1. A total of 167 eligible CRT subjects were identified, of whom 105 (63%) were V-paced>40% at final follow-up prior to commencement of CRT. 122 (73%) had congenital heart disease, the majority of whom (85 (70%)) had a biventricular circulation with systemic LV. A structural surgical procedure was performed within 30days before or after the implant of the CRT system in 34 (20%) subjects, 29 (85%) of which were at the same procedure.

**Table 1.**
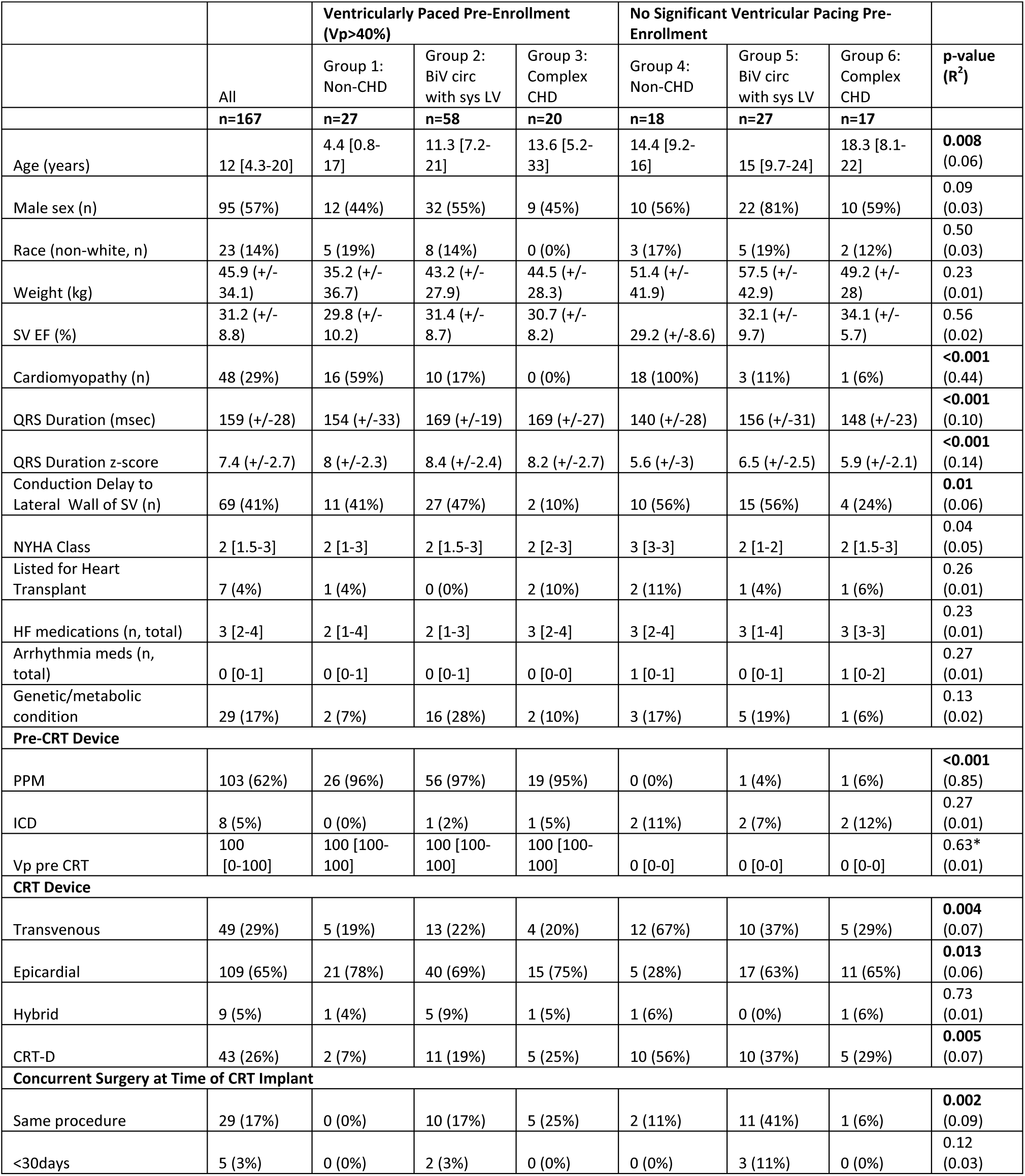
Baseline Demographics. BiV: biventricular, SV EF: systemic ventricle ejection fraction, NYHA: New York Heart Association, HTX: heart transplant listing prior to implant, HF: heart failure, PPM: permanent pacemaker, Vp: ventricular pacing, CRT-D: cardiac resynchronization therapy-defibrillator.

Between the 6 pre-defined subgroups, several trends in baseline indices were noted. Those with prior pacing (Groups 1, 2 and 3) had a very high median pacing percentage (100% [100- 100%]). Compared to the non-paced cohorts (Groups 4,5 and 6) they were similar in age at enrollment (9.8 [3.8-20] years versus 14.9 [9.2-20] years, p=0.13) and had similar SVEF (30.8±8.9% versus 31.8±8.5%, p=0.49), but tended to have wider QRSd at baseline (165±26ms versus 149±28ms, p=0.0003).

CRT delivery was good throughout follow-up with median pacing percentage 100% [100-100%] at all follow-up time points. Follow-up was recorded close to the target dates of 6months post implant (actual timing 5.8months [4.1-6.7months] post implant), 12 months (12.4months [11.1- 13.9months]), 5 years (5.0 [4.7-5.4years]) and latest follow-up (5.1 years [2.0-8.6 years]).

### Response Definition 1: Improvement in SVEF within the First Year Post-CRT

#### Response to CRT

Formal assessment of SVEF was available for 150 (90%) subjects for at least one of the stipulated 6 month or 12 month time points. 90 (60%) had follow-up available at both time points, with excellent intraclass correlations between the SVEF measurements made at the two time points (average ICCs, two-way mixed effects, absolute agreement: SVEF 0.91 (95% CI 0.86- 0.94)). Average values were taken for those with two follow-up measurements.

Across all subjects with formal SVEF measurements, 96 (64%) exhibited an improvement in absolute SVEF >5% and 110 (73%) a proportional improvement in SVEF >10% (Table 2 and Figure 2). Overall, there was an median increase in SVEF of 11% [3-21%] (p<0.001 compared to baseline), and a decrease in QRS duration (18±24msec, p<0.001), QRSd z-score (1.9±2.4, p<0.001) and NYHA class (0.6±1 lower, p<0.001), but no change in number of HF medications (0.04±0.92, p=0.63). Between subgroups, there was minimal variation in degree of change of clinical parameters, except for QRSd and QRSd z-score, which shortened more in the prior paced groups (delta of 22±21ms versus 8±25ms, p<0.001 and delta of 2.5±2.1 versus 1.0±2.5, p<0.001, respectively).

**Figure 2.**
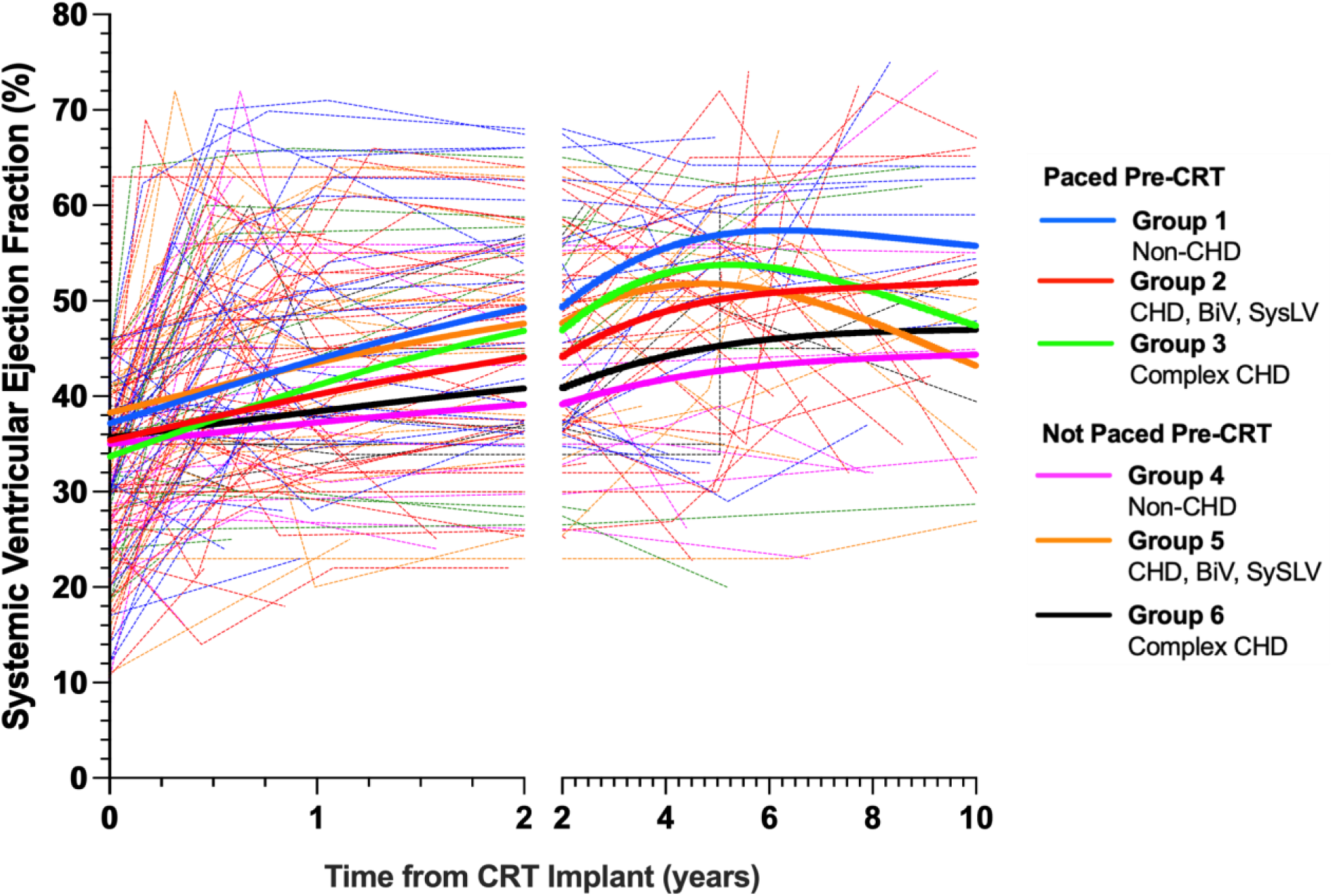
Change in systemic ventricular ejection fraction with time from CRT implant (split x- axis). CHD: congenital heart disease, BiV: biventricular, SysLV: systemic left ventricle. Best fit line calculated using cubic regression.

**Table 2.**
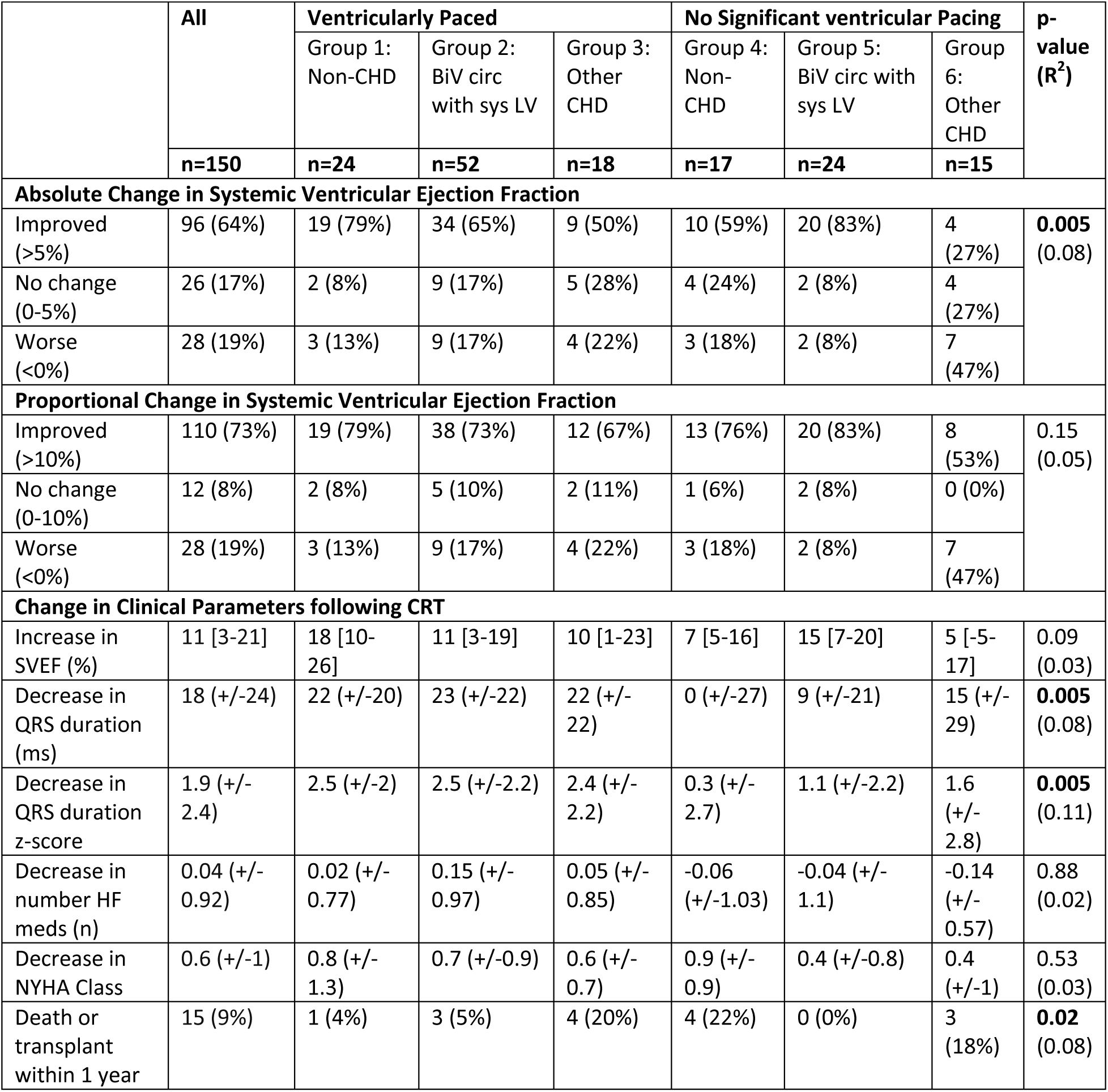
Response to CRT by Subgroup. Please see methods for definitions of improvement/no change/worsening of primary Endpoints. p-value is one-way ANOVA, grouped by the six subtypes.

|  | All | Ventricularly Paced |  |  | No Significant ventricular Pacing |  |  | p-value<br>(R<sup>2</sup>) |
| --- | --- | --- | --- | --- | --- | --- | --- | --- |
|  |  | Group 1:<br>Non-CHD | Group 2:<br>BiV circ<br>with sys LV | Group 3:<br>Other<br>CHD | Group 4:<br>Non-CHD | Group 5:<br>BiV circ with<br>sys LV | Group 6:<br>Other<br>CHD |  |
|  | n=150 | n=24 | n=52 | n=18 | n=17 | n=24 | n=15 |  |
| Absolute Change in Systemic Ventricular Ejection Fraction |  |  |  |  |  |  |  |  |
| Improved (>5%) | 96 (64%) | 19 (79%) | 34 (65%) | 9 (50%) | 10 (59%) | 20 (83%) | 4 (27%) | 0.005<br>(0.08) |
| No change (0-5%) | 26 (17%) | 2 (8%) | 9 (17%) | 5 (28%) | 4 (24%) | 2 (8%) | 4 (27%) |  |
| Worse (<0%) | 28 (19%) | 3 (13%) | 9 (17%) | 4 (22%) | 3 (18%) | 2 (8%) | 7 (47%) |  |
| Proportional Change in Systemic Ventricular Ejection Fraction |  |  |  |  |  |  |  |  |
| Improved (>10%) | 110 (73%) | 19 (79%) | 38 (73%) | 12 (67%) | 13 (76%) | 20 (83%) | 8 (53%) | 0.15<br>(0.05) |
| No change (0-10%) | 12 (8%) | 2 (8%) | 5 (10%) | 2 (11%) | 1 (6%) | 2 (8%) | 0 (0%) |  |
| Worse (<0%) | 28 (19%) | 3 (13%) | 9 (17%) | 4 (22%) | 3 (18%) | 2 (8%) | 7 (47%) |  |
| Change in Clinical Parameters following CRT |  |  |  |  |  |  |  |  |
| Increase in SVEF (%) | 11 [3-21] | 18 [10-26] | 11 [3-19] | 10 [1-23] | 7 [5-16] | 15 [7-20] | 5 [-5-17] | 0.09<br>(0.03) |
| Decrease in QRS duration (ms) | 18 (+/-24) | 22 (+/-20) | 23 (+/-22) | 22 (+/-22) | 0 (+/-27) | 9 (+/-21) | 15 (+/-29) | 0.005<br>(0.08) |
| Decrease in QRS duration z-score | 1.9 (+/-2.4) | 2.5 (+/-2) | 2.5 (+/-2.2) | 2.4 (+/-2.2) | 0.3 (+/-2.7) | 1.1 (+/-2.2) | 1.6 (+/-2.8) | 0.005<br>(0.11) |
| Decrease in number HF meds (n) | 0.04 (+/-0.92) | 0.02 (+/-0.77) | 0.15 (+/-0.97) | 0.05 (+/-0.85) | -0.06 (+/-1.03) | -0.04 (+/-1.1) | -0.14 (+/-0.57) | 0.88<br>(0.02) |
| Decrease in NYHA Class | 0.6 (+/-1) | 0.8 (+/-1.3) | 0.7 (+/-0.9) | 0.6 (+/-0.7) | 0.9 (+/-0.9) | 0.4 (+/-0.8) | 0.4 (+/-1) | 0.53<br>(0.03) |
| Death or transplant within 1 year | 15 (9%) | 1 (4%) | 3 (5%) | 4 (20%) | 4 (22%) | 0 (0%) | 3 (18%) | 0.02<br>(0.08) |

There was minimal correlation between change in EF and change in QRSd (p=0.10, R^2^=0.01) or QRSd z-score (p=0.044, R^2^= 0.03), suggesting that the crude shortening of QRSd was not highly significant in determining EF improvement following CRT. Improvement in EF was more strongly correlated with improvement in NYHA class (p<0.001, R^2^=0.12).

#### Morphological and pathophysiological predictors of response

Factors associated with the response to CRT by ordinal logistic regression are shown in Table 3. The pathophysiological groups 1-5 were not significantly associated with response, and only Group 6 (no prior pacing, complex CHD) differed significantly from the other groups. This group was least likely to demonstrate a response to CRT in terms of either an absolute (coefficient - 2.2 [-3.6 --0.8), p=0.002) or proportional response (coefficient-1.4 [-2.9 --0.03), p=0.046).

**Table 3.** Association of baseline characteristics and change in systemic ventricular ejection fraction following CRT. Ordinal logistic regression, with categories as defined in methods. * For the multivariable analysis of Subgroup 6, Systemic LV and biventricular circulation were excluded due to collinearity.

|  | Absolute Change in Systemic Ventricular Ejection Fraction |  |  |  | Proportional Change in Systemic Ventricular Ejection Fraction |  |  |  |
| --- | --- | --- | --- | --- | --- | --- | --- | --- |
|  | Univariable |  | Multivariable |  | Univariable |  | Multivariable |  |
|  | Coef | p-value | Coef | p-value | Coef | p-value | Coef | p-value |
| Age (years) | -0.02 (-0.04-0) | 0.062 |  |  | -0.02 (-0.04-0) | 0.041 | -0.01 (-0.03-0.02) | 0.567 |
| Sex (male) | -0.33 (-0.99-0.32) | 0.32 |  |  | -0.81 (-1.58--0.05) | 0.038 | -0.64 (-1.51-0.24) | 0.153 |
| Race (non-white) | 0.38 (-0.7-1.46) | 0.49 |  |  | -0.02 (-1.11-1.06) | 0.97 |  |  |
| Weight (kg) | -0.01 (-0.02-0) | 0.2 |  |  | -0.01 (-0.02-0) | 0.25 |  |  |
| Systemic Ventricle EF (%) | -0.05 (-0.09--0.01) | 0.013 | -0.05 (-0.09-0) | **0.039** | -0.08 (-0.12--0.03) | 0.001 | -0.08 (-0.13--0.02) | **0.006** |
| Cardiomyopathy (n) | -0.22 (-0.91-0.48) | 0.55 |  |  | -0.39 (-1.14-0.36) | 0.31 |  |  |
| Biventricular Circulation | 1.07 (0.15-1.99) | 0.022 | 0.86 (-0.16-1.88) | 0.1 | 0.53 (-0.53-1.59) | 0.33 |  |  |
| Systemic LV | 0.96 (0.14-1.78) | 0.021 | 0.4 (-0.56-1.35) | 0.417 | 0.64 (-0.27-1.55) | 0.17 |  |  |
| Congenital Heart Disease | -0.41 (-1.17-0.35) | 0.29 |  |  | -0.35 (-1.2-0.49) | 0.41 |  |  |
| Prior pacing (Vp>40%) | 0.24 (-0.43-0.91) | 0.48 |  |  | 0.07 (-0.67-0.81) | 0.86 |  |  |
| QRS duration (ms) | -0.01 (-0.02-0) | 0.11 |  |  | -0.02 (-0.03-0) | 0.037 | -0.02 (-0.03-0) | 0.061 |
| QRS duration z-score | -0.04 (-0.17-0.08) | 0.51 |  |  | -0.06 (-0.2-0.08) | 0.41 |  |  |
| Conduction delay to Lat Wall SV | 0.94 (0.23-1.66) | 0.01 | 0.7 (-0.03-1.50) | 0.059 | 1.21 (0.36-2.07) | 0.006 | 1.28 (0.36-2.2) | **0.006** |
| Lead Separation Ratio | -3.47 (-6.86--0.09) | 0.044 | -3.06 (-6.66-0.55) | 0.097 | -3.65 (-7.34-0.04) | 0.053 |  |  |
| NYHA Class | -0.11 (-0.49-0.27) | 0.58 |  |  | -0.04 (-0.47-0.38) | 0.84 |  |  |
| Number of HF Medications (n) | -0.11 (-0.35-0.13) | 0.37 |  |  | -0.01 (-0.28-0.26) | 0.94 |  |  |
| Genetic/Metabolic Disorder | 0.41 (-0.48-1.29) | 0.37 |  |  | -0.02 (-0.92-0.87) | 0.96 |  |  |
| Subtype 1 | NA | NA |  |  | NA | NA |  |  |
| Subtype 2 | -0.67 (-1.8-0.47) | 0.25 |  |  | -0.34 (-1.49-0.81) | 0.56 |  |  |
| Subtype 3 | -1.21 (-2.51-0.1) | 0.071 |  |  | -0.64 (-2.01-0.73) | 0.36 |  |  |
| Subtype 4 | -0.89 (-2.24-0.47) | 0.2 |  |  | -0.2 (-1.68-1.29) | 0.8 |  |  |
| Subtype 5 | 0.3 (-1.15-1.76) | 0.68 |  |  | 0.29 (-1.16-1.73) | 0.7 |  |  |
| Subtype 6 | -2.2 (-3.57--0.83) | 0.002 | -1.4 (-2.4 - -0.4)* | **0.008** * | -1.44 (-2.86--0.03) | 0.046 | -6.5 (-2.5--0.2)<sup>†</sup> | **0.021** |

Other predictors of response differed between the absolute and proportional SVEF improvement assessments. On analysis of predictors of absolute SVEF improvement (>5% rise in SVEF), patients with lower baseline SVEF were more likely to experience a beneficial response, on both univariable and multivariable analysis. Those with biventricular circulation, systemic LV, CDtoLWSV and lower lead separation ratio were more likely to respond on univariable but not multivariable analysis.

On assessment of predictors of proportional improvement in SVEF, a lower SVEF was again a significant predictor on univariable and multivariable response, as was CDtoLWSV.

The association of SVEF and response (absolute and proportional) is demonstrated in Figure 3. The coefficient for response was significantly worse for those with higher SVEF. All higher SVEF groups were associated with significantly lower response coefficient (p<0.05) on absolute response, with the exception of the 3^rd^ tertile (36/45%) and 4^th^ quintile (36/40%). All higher SVEF groups were associated with significantly lower response coefficient on proportional response, with the exception of the 2^nd^ quartile (25/31%).

**Figure 3.**
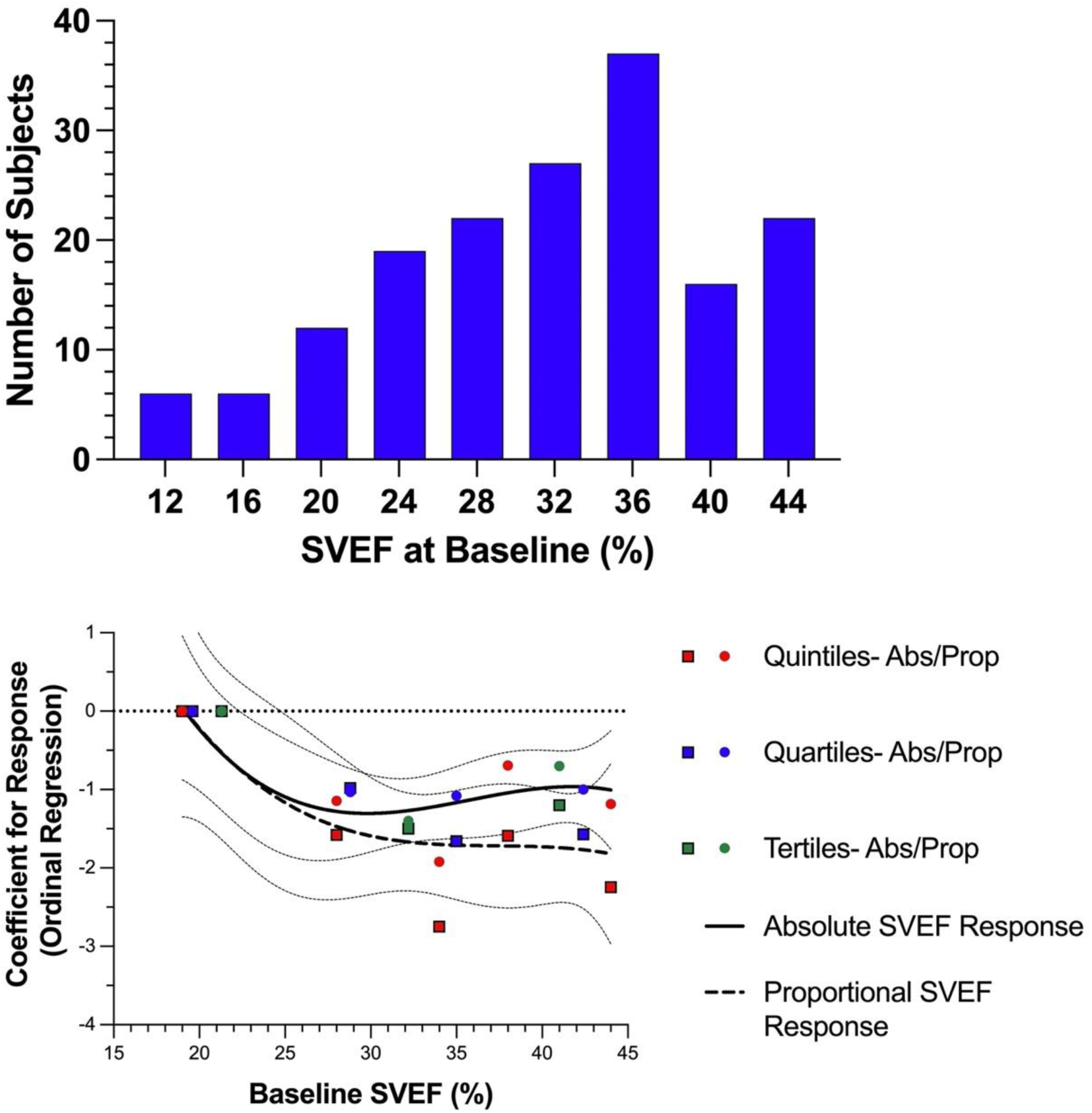
(Upper) Frequency histogram of systemic ventricle ejection fraction (SVEF) at baseline. (Lower) Coefficient for response (ordinal regression) versus lowest quintile/quartile/tertile for absolute SVEF response (Abs) and proportional SVEF response (Prop). Upper and lower tertile cutoffs were 10/28%, 28/35% and 36/45%, respectively. Quartile cutoffs were 10/25%, 25/31%, 31/37% and 37/45%. Quintile cutoffs were 10/24%, 24/30%, 30/35%, 36/40%, and 40/45%.

### Response Definition 2: Longitudinal Improvement in Parameters versus Control Groups

#### Response to CRT

Across the whole cohort (139 propensity matched pairs, total 278 subjects), there was a clear longitudinal impact of CRT upon SVEF (CRT coefficient 4.0 (1.9-6.1), p<0.001) and QRS duration z-score (-0.7 (-1.2--0.2), p=0.015). There was no significant CRT-associated longitudinal impact upon NYHA class, total number of HF medications, or somatic growth (in terms of weight or height). There remained no association of CRT with somatic growth on subanalysis of subjects prior to age 18years (192 PSM subjects, Height: 493 measurements, CRT coefficient 1.02 (-8.7- 11), p=0.84); Weight: 507 measurements, CRT coefficient 0.95 (-6.4-8.3), p=0.80).

#### Morphological and pathophysiological predictors of response

Longitudinal change in SVEF was assessed within the primary binary subgroups [with/without pacing, with/without CHD, with/without biventricular circulation, with systemic LV or RV]. A significant positive association of SVEF with CRT was seen in 6 of the 8 subgroups (75%) (Figure 4). The relationship between CRT and longitudinal SVEF was not significant in patients with a systemic RV, those who were not paced prior to enrollment, and in subgroups with small numbers.

**Figure 4.**
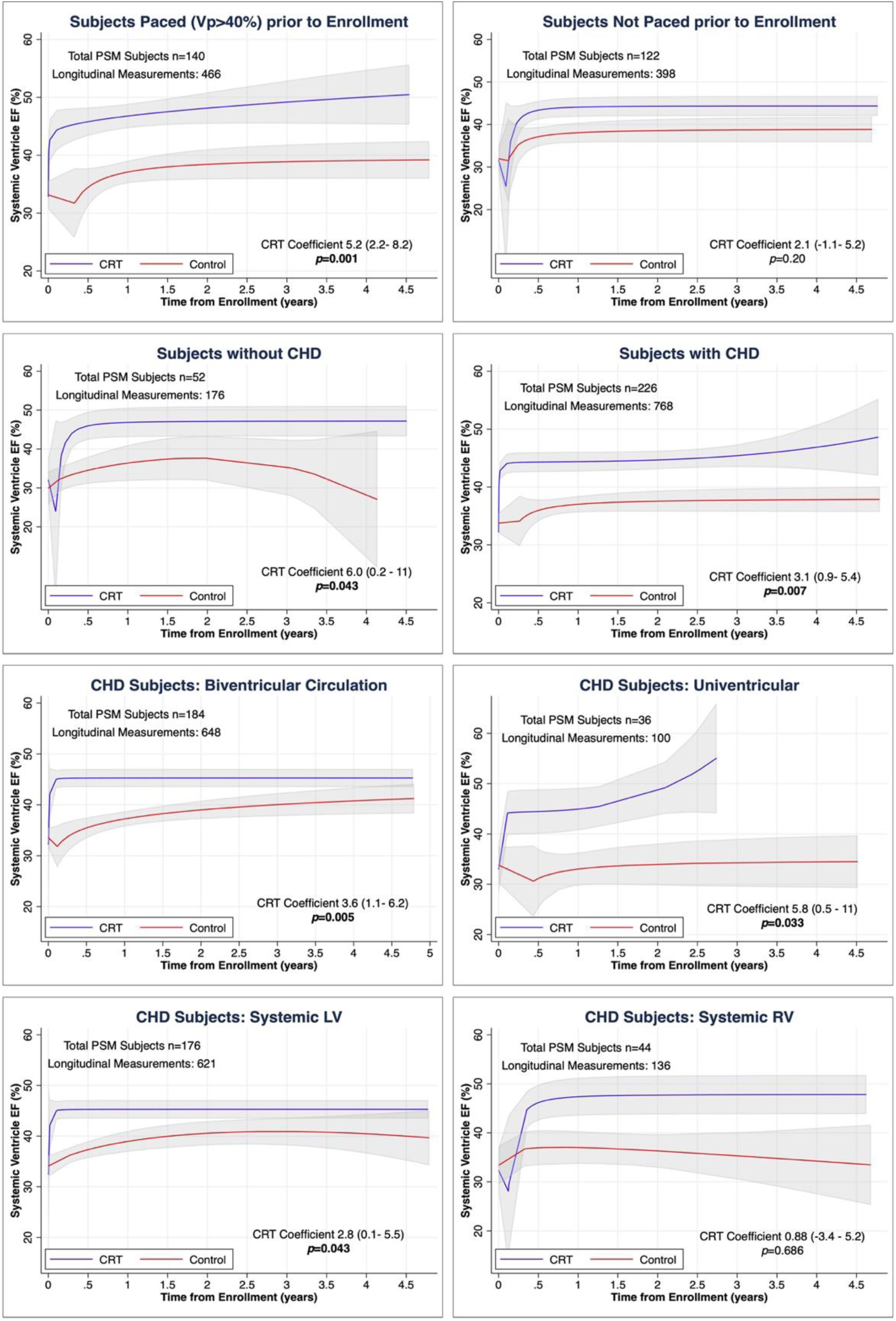
Longitudinal plots presenting the long-term trajectory of systemic ventricle ejection fraction (SVEF) within the eligible propensity score matched (PSM) cohorts with and without CRT. Separate PSM was performed for each subanalysis. Plots are fractional polynomial plots and the pacing effect is estimated using multilevel mixed-effects linear regression modeling with cubic model.

### Sensitivity analyses

Excluding subjects who underwent concurrent surgical procedures within 30days either side of CRT implant (n=34), overall change in SVEF was similar at +10% [3-21%] across all 118 subjects with early formal SVEF assessment. 74 (63%) were responders in terms of absolute improvement in SVEF and 87 (74%) were responders in terms of proportional improvement in SVEF. Significant univariable and multivariable predictors of response remained unchanged, except that CDtoLWSV was now a significant predictor of absolute response on multivariable analysis (0.93 [0.06-1.83], p=0.032) as well as univariable. On longitudinal analysis of PSM subjects and controls (252 PSM subjects, 830 measurements), SVEF remained significantly associated with CRT (CRT coefficient 2.7 (0.2-5.2), p=0.038).

## Discussion

This study assessed the response to CRT in a large multicenter retrospective cohort of children and patients with CHD. Two measures of response were assessed. One measure used the CRT recipient as their own control, defining response as an improvement in SVEF in the medium term at 6-12months. The second measure used a tightly matched control cohort of non-CRT patients, and compared the longitudinal trajectories of the patients with and without CRT. The main findings are:

- Medium-term improvement in SVEF from baseline

⚬ A majority of subjects responded to CRT, with responders identified within all subgroups
⚬ Those with low baseline EF or conduction delay to the lateral wall of the systemic ventricle (for example, LBBB in a structurally normal heart) were most likely to respond.
⚬ Those without prior ventricular pacing <u>and</u> with ‘complex’ CHD were less likely to respond
⚬ QRS duration was not associated with response
- Improved long-term trajectory versus controls

⚬ CRT was associated with a general improvement in SVEF and shortening of QRS duration
⚬ Most binarised subgroups demonstrated an improvement in SVEF with CRT
⚬ Those with systemic RV and those without pacing prior to enrollment did not demonstrate a significantly improved SVEF trajectory compared to controls

### Patient Anatomical and Pathological Variants

By the nature of the patient cohort, there was considerable heterogeneity between each subject. Each subject presents a unique constellation of patient-specific factors, all of which may impact upon CRT response. There is a constant pull between the desire to define the patient groups as homogenously as possible versus the obligation to avoid creating ever smaller divisions that preclude the possibility of meaningful comparison. The six groups were designed to map onto those patient groups that have been defined in prior studies to be potentially good or poor responders to CRT.^11^ These include pacing-induced cardiomyopathies, systemic LVs, systemic RVs and univentricular circulations ^7–9, 20, 21^.

### Highest Likelihood of Response

The absence of consistent significant associations between most of the anatomical and pathological groups and subsequent response to CRT is a significant finding. On assessment of early response (using the patient as their own baseline), the most consistent finding was that patients with lower SVEF tended to be most likely to demonstrate not only a proportional response, but also an absolute response (>5% improvement). This suggests that there may not be a clear cutoff to define when CRT is ‘too late’, and that even those with severely impaired function may experience an increase in SVEF with CRT.

Conduction delay to the lateral wall of the systemic ventricle was also a parameter that was significant on univariable analysis of absolute response, and both univariable and multivariable analysis of proportional response. This measure was selected as analogous to LBBB in the functionally normal heart, and may be an important indicator in more complex hearts. Further research is required to assess whether this is associated with epiphenomena such as classical pattern dyssynchrony in the CHD population.^22^ However, the fact that it was rarely seen in the complex CHD groups (16% of subjects), and yet the response rate in those groups was 40-60%, suggests that this relatively crude index of electrical activation is a useful but not an essential criterion for response to CRT.

In contrast to many non-CHD adult CRT studies, it was notable that increased QRSd alone was not associated with response by any measure. Some of this difference may be accounted for by the heterogeneity of interventricular conduction delays, but the most significant fact is likely that the vast majority of recipients had a ‘sufficient’ degree of electrical dyssynchrony. 156 (93%) had a QRSd z-score>3.55 (the cutoff employed as a marker of equivalency for strict LBBB criteria), and 145 (87%) had QRSd z-score >4.8, the equivalent to around 150msec and the longest QRSd cutoff evaluated in major conventional CRT trials.

### Lower Likelihood of Response

On longitudinal assessment versus controls, patients without prior ventricular pacing and those with systemic RV did not demonstrate a superior outcome with CRT (Figure 4). This trend was also reflected in the lower medium-term response rates in the group with no prior pacing, and CHD without a biventricular, systemic LV, circulation. Again, in such a heterogenous group, identifying the underlying reasons for these findings is challenging, but a number of factors may be postulated.

#### CRT in Patients without Biventricular Circulation

Increasingly, large studies have demonstrated that ventricular pacing of the single ventricle patient is highly deleterious to long term survival, but no study has identified a clear improvement in outcome with CRT.^23, 24^ At least in part, this is likely related to the limited number of single ventricle patients receiving CRT systems. Numbers in this study remain relatively small, but nevertheless, we identified a significant impact of CRT on long-term SVEF trends. The second response outcome of the study, longitudinal trends versus matched controls, assists determining whether differing responses to CRT are related to weaker effect of CRT or the higher overall risk of deterioration in specific patient groups. In this context, there is an argument that, for patients with a single ventricle, the absence of deterioration may itself be a good response to CRT, and this would explain the discrepancy between the univariable findings of medium-term response (where there is a trend to better CRT response in biventricular patients) and the longitudinal response versus controls. A similar trend in the single ventricle population has been demonstrated previously by the Boston group.^25^

#### CRT in Patients with Systemic Right Ventricle

The other major anatomical subgroup that has received individual attention in most prior studies is patients with systemic RV. Published results in this cohort have been mixed at best. While some studies have found an attenuated response to CRT ^8, 9, 26^, others showed no response.^7, 21^ A recent metanalysis found a marginally inferior response in the systemic RV population.^11^ Our study again found a mixed picture. On medium-term SVEF response, there was a weak (univariable only) trend towards poorer response to CRT, while on longitudinal outcome versus controls there was no significant CRT effect. These findings may be partially attributed to challenges in measuring the EF of the systemic RV, and the latter finding may relate predominantly to sample size (Figure 4). As such, these findings certainly do not suggest that CRT is contraindicated in the systemic RV population, but there is at least weaker evidence of efficacy.

### Clinical Implications

This study has demonstrated that CRT results in a clear improvement in SVEF in many pediatric and CHD patients, with responders identified in all anatomical and pathophysiological subtypes.

However, CRT remains a complex intervention with significant risks anticipated both at the time of implant (particularly in those requiring repeat sternotomy) and in a life-time of device management. Therefore, indices to identify those who are most likely to respond remain helpful in weighing risk and benefit. On assessment of both medium-term change in SVEF and longitudinal trajectory versus controls there are significant trends that should be noted. Based upon this data, those with lower SVEF, those with electrical conduction delay to the lateral wall of the systemic ventricle (such as LBBB in those with conventional ventricular orientation) and those with systemic LV are most likely to experience a medium and long-term response.

### Limitations

This was a multicenter retrospective study, and as such the delineation of a clinical response to CRT is highly challenging. We have attempted to be as comprehensive as possible, using two outcome measures, one indexed to the patient and one to a tightly matched control group. It is also established that SVEF alone is not the only marker of response, and prospective studies would be significantly enhanced by the formal evaluation of baseline parameters such as cardiopulmonary metabolics and laboratory studies such as BNP or NT-proBNP.

We chose to include patients with concurrent surgery in the main analyses. Many of these surgical procedures were on the subpulmonary ventricle, but nonetheless, there are inevitably hemodynamic implications for the systemic ventricle and general clinical condition resulting from all structural surgical interventions. Sensitivity analyses suggested a similar result for the main clinical analyses.

In a similar vein, in common with all major studies of CRT in these patient cohorts, the pediatric and CHD groups have been amalgamated for many of the analyses. HF in pediatric patients without CHD represents a very different pathology to HF in those with CHD, and there is an argument that they should be assessed entirely separately. However, the numbers of patients in each subgroup were small and we have generally aimed for a robust assessment on multivariable analysis to assess and quantify the role of contributing factors.

## Conclusions

CRT in children and patients with CHD frequently results in an improvement in SVEF. This study suggests that all the major anatomical and pathophysiological subgroups have the potential to respond to CRT. However, those with lower SVEF, those with electrical conduction delay to the lateral wall of the systemic ventricle (such as LBBB in those with conventional ventricular orientation), and those with systemic LV are most likely to experience a medium-and long-term response.

## Abbreviations List

CHD: Congenital Heart Disease
CRT: Cardiac Resynchronization Therapy
DCM: Dilated Cardiomyopathy
EF: Ejection Fraction
HF: Heart Failure
HR: Hazard Ratio
ICD: Implantable Cardioverter Defibrillator
LV: Left Ventricle
NYHA: New York Heart Association
PSM: Propensity Score-Match
QRSd: QRS Duration
RV: Right Ventricle
SV: Systemic Ventricle

## Data Availability

Anonymized data is available on reasonable request following publication of the manuscript

